# Rapid surveillance platforms for key SARS-CoV-2 mutations in Denmark

**DOI:** 10.1101/2021.10.25.21265484

**Authors:** Katja Spiess, Vithiagaran Gunalan, Ellinor Marving, Sofie Holdflod Nielsen, Michelle G. P. Jørgensen, Anna S. Fomsgaard, Line Nielsen, Alonzo Alfaro-Núñez, Søren M. Karst, The Danish COVID-19 Genome Consortium (DCGC), Shila Mortensen, Morten Rasmussen, Ria Lassaunière, Maiken Worsøe Rosenstierne, Charlotta Polacek, Jannik Fonager, Arieh S. Cohen, Claus Nielsen, Anders Fomsgaard

**Author notes:** https://www.covid19genomics.dk/about (The Danish COVID-19 Genome Consortium (DCGC)). These authors contributed equally (Katja Spiess, Vithiagaran Gunalan). Corresponding author: Katja Spiess.

## Abstract

Multiple mutations in SARS-CoV-2 variants of concern (VOCs) may increase, transmission, disease severity, immune evasion and facilitate zoonotic or anthoprozoonotic infections. Four such mutations, ΔH69/V70, L452R, E484K and N501Y, occur in the SARS-CoV-2 spike glycoprotein in combinations that allow detection of the most important VOCs. Here we present two flexible RT-qPCR platforms for small-and large-scale screening to detect these mutations, and schemes for adapting the platforms for future mutations. The large-scale RT-qPCR platform, was validated by pair-wise matching of RT-qPCR results with WGS consensus genomes, showing high specificity and sensitivity. Detection of mutations using this platform served as an important interventive measure for the Danish public health system to delay the emergence of VOCs and to gain time for vaccine administration. Both platforms are valuable tools for WGS-lean laboratories, as well for complementing WGS to support rapid control of local transmission chains worldwide.

## Introduction

The global SARS-CoV-2 pandemic, which raised with the identification of this novel coronavirus in late 2019, has seen the emergence of several variants, each with a distinct set of mutations^1^. Early detection of new SARS-CoV-2 mutations and associated measures to decrease the risk of spread are important to control local outbreaks of SARS-CoV-2 variants, especially those which have been designated Variants of Concern (VOCs)^2,3^. The latter are defined by increased transmissibility, severity of infections and resistance to immunity^4–8^. VOCs include the Alpha (B.1.1.7) and (B.1.1.7 + E484K), Beta (B.1.351), Gamma (P1) and Delta (B.1.617.2) variants (**Fig.1A-C/Tab. 1**).

**Figure 1.**
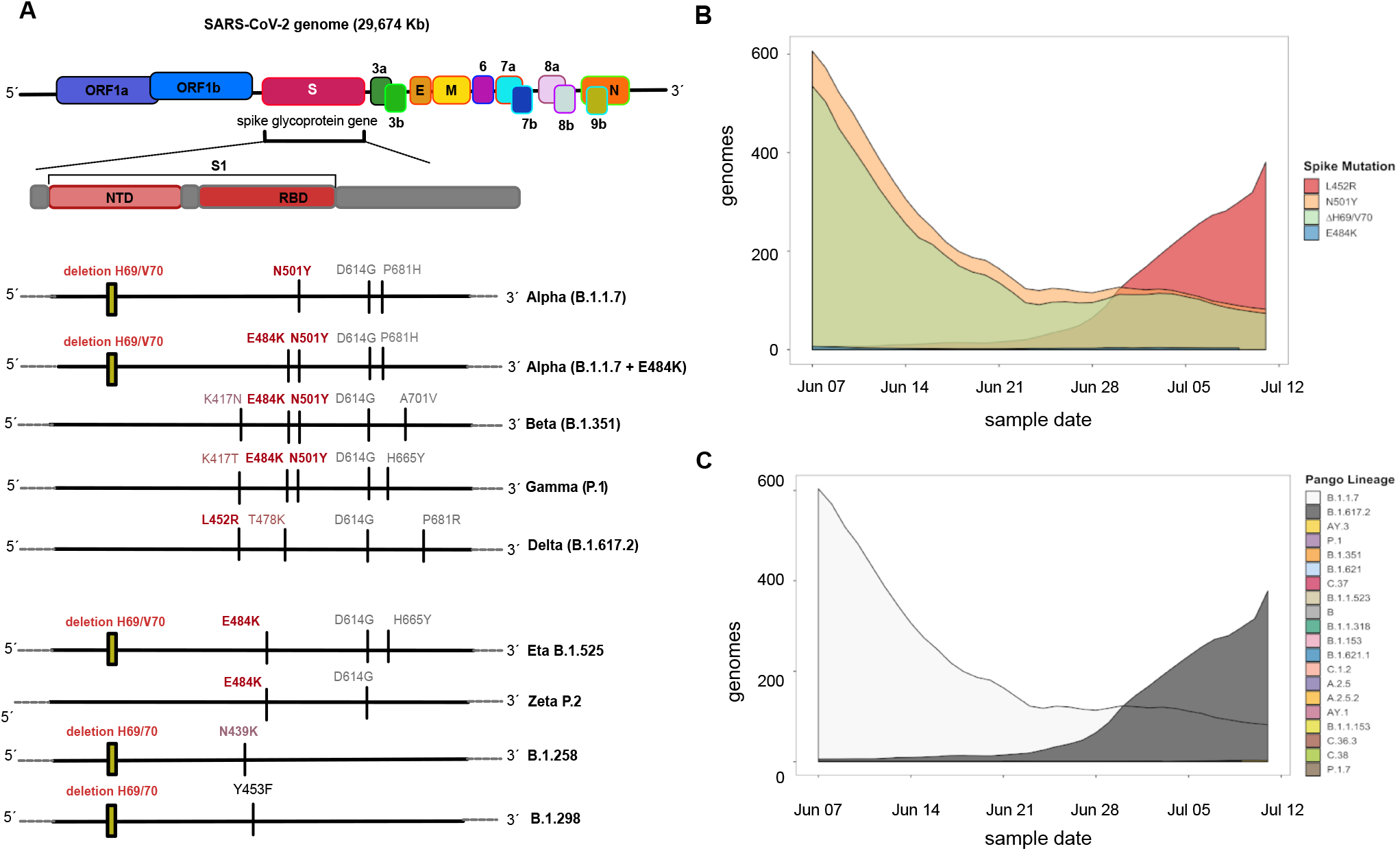
Overview of the key mutations located in the spike glycoprotein during the pandemic and PCR strategies. **(A)** The spike glycoprotein is located between the ORF1B and 2a within the SARS-CoV-2 genome. The ΔH69/V70 (2 amino acid deletion) is located in the N-terminal domain (NTD) of the spike glycoprotein and the L452R, E484K and N501Y mutations are located in the receptor-binding domain (RBD). Sets of four variant specific mutations present in VOC. The Beta (B.1.351) and the Gamma (P.1) have the same key mutations. The y mutations are also present SARS-CoV2 variants that are not variants of concern, but the variants are included into this study for detecting the mutations in patient samples. **B)** Prevalence of spike mutations ∆H69/V70, N501Y, E484K, L452R amongst SARS-CoV-2 consensus genomes in Denmark between 7^th^ of June 2021 to the 11^th^ of July 2021 (**C)** Variant composition (by Pangolin nomenclature) harbouring key spike mutations.

**Table 1.**
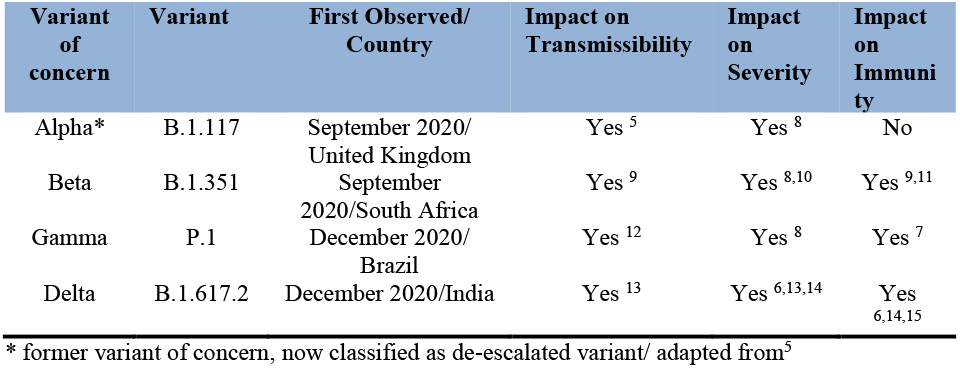
Overview of SARS-CoV-2 variants, occurrence and evidence of impact.

In all these VOCs, combinations of key mutations are present in *S*: N501Y in the variants Alpha (B.1.1.7), Beta (B.1.351) and Gamma (P.1); E484K in the variants Beta (B.1.351) and Gamma (P.1) and within the emerging Alpha B.1.1.7 variant^16^; L452R in the Delta (B.1.617.2) variant and ΔH69/V70 in the variants Alpha (B.1.1.7) and B.1.1.298.

The N501Y mutation occurs in the receptor-binding interface and confers a substantial increase in the binding affinity of the *S* for the human angiotensin-converting enzyme 2 (hACE2) protein^17^. HACE2 interaction with the *S* is essential for virus entry and infection of the cells^18^. The E484K mutation has been identified as an immunodominant spike protein residue, facilitating escape from several monoclonal antibodies, as well as antibodies in convalescent plasma^19–21^. Altered immune recognition has also been described for the L452R mutation^21–23^. The key mutation ΔH69/V70, a two amino acid deletion, has appeared in multiple SARS-CoV-2 variants at different geographical locations across Europe. In Denmark, ΔH69/V70 was detected in local outbreaks in mink farms in Northern Jutland ^24,25^. The spread of this deletion in combination with additional mutations (notably Y453F) resulted in the SARS-CoV-2 mink variants B.1.1.298, which transmitted both ways between humans and mink; also giving rise to the early “cluster 5” variant ^24,26.^.

The identification of these variants and the mutations that form their signature are largely dependent on Whole Genome Sequencing (WGS) of SARS-CoV-2 from infected individuals. In addition, WGS of SARS-CoV-2 also elucidates sets of novel mutations potentially linked to changes in viral properties or associated with vaccine breakthrough. However, the utility of WGS in a pandemic such as this also carries with it a significant cost in the form of reagents, equipment as well as turnaround time – the average time from sample to genome being ∼1-7 days depending on the scale of sequencing performed. This has led to the development of alternatives to WGS such as SARSeq, which is based on sequencing of the ectodomain of the SARS-CoV-2 spike protein^27^ or sequencing of the whole S gene using Sanger sequencing^28^.

While such approaches yield cost and reagent savings, the turnaround time, preparation effort for these and cost are still higher compared to RT-qPCR detection platforms. In addition, qPCR technology is inarguably one of the cornerstones of modern infectious disease diagnostics, thus expertise and equipment is readily available and is not hindered by technical issues that might present themselves with newer technologies, which could potentially delay the implementation of such screening approaches. In order to detect SARS-CoV-2 mutations in near real-time after sample acquisition and to allow for implementation at different scales (both small and large), we developed fast, robust and flexible RT-qPCRs platforms using state-of-the-art modified detection probes. Small-scale screening entails the simultaneous detection of three key mutations in a multiplexed RT-qPCR, where sets of variant-specific mutations can be replaced by a single signature mutation of concern such as for the Delta (B.1.617.2) variant with the L452R mutation. The large-scale screening strategy entails the detection of four key mutations by a combination of multiplexed and single RT-qPCRs running in parallel in a 384-well format.

Validation of the large-scale implementation of this RT-qPCR platform was performed for 9572 positive samples collected between 6^th^ June 2021 and 11^th^ July 2021 as part of the national surveillance program in Denmark using paired WGS consensus genomes derived from SARS-CoV-2 positive samples. From here, the specificity, sensitivity, Positive Predictive Value (PPV) and Negative Predictive Value (NPV) were determined for the large-scale RT-qPCR platform. The RT-qPCR platforms for both small- and large-scale screening are designed as flexible detection systems, where new mutations of concern can be included, thereby following the course of the pandemic with minimal lag time.

## RESULTS

### SMALL SCALE SCREENING OF SARS-COV-2 VARIANTS OF CONCERN

For laboratories with small amounts of positive SARS-CoV-2 samples or without the capacity to screen on a large scale for SARS-CoV-2 variants we developed a multiplexed RT-qPCR (**v.1**) that can detect three key mutations (ΔH69/V70, E484K and N501Y) simultaneously (**Fig. 2A**). As proof of concept to determine if a key mutation can be replaced by another, we replaced the ΔH69/V70 with the L452R mutation present in the delta variant (B.1.617.2) in the multiplex RT-qPCR (**v.2**) (**Fig. 2B)**.

**Figure 2.**
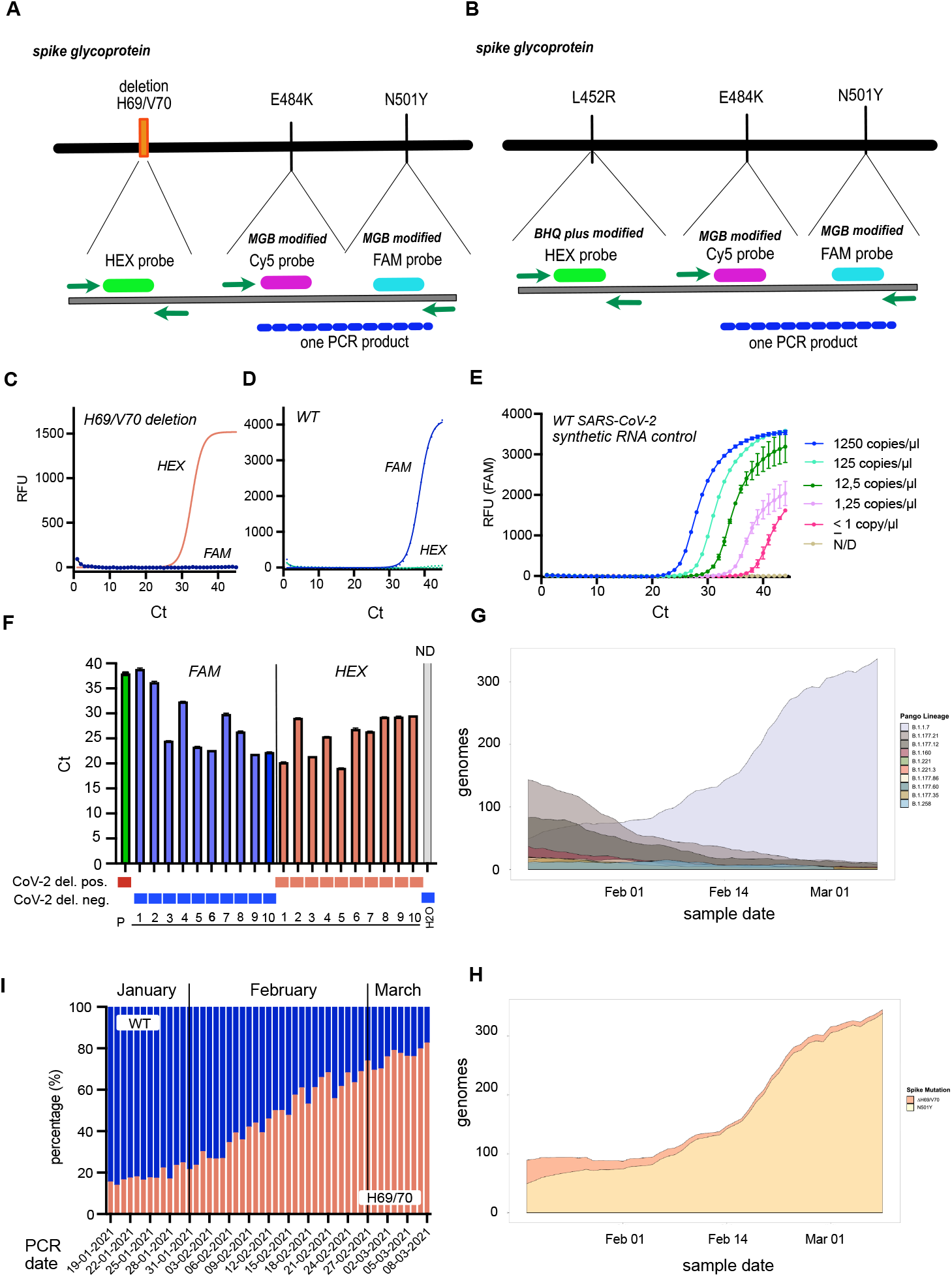
Schematic overview about the PCR platforms and establishment of the H69/70 RT-qPCR. **(A)** Multiplexed RT-qPCR v.1 targeting ∆H69/70V, E484K- and N501Y mutation. The deletion and mutations are detected with one probe respectively and the E484K and N501Y mutations are detected by one primer pair resulting in a single amplification product for both mutations. (**B)** As proof of concept ∆H69/70V was replaced by the L452R mutation of the delta variant (B.1.617.2) in the multiplexed RT-qPCR v.2. (**C)** A HEX-labelled probe detects the ΔH69/70 and (**D)** a FAM-labelled probe the WT nucleotide sequence. (**E)** Dilution row of the TWIST control (WT SARS-CoV-2) to detect the limit of detection (**F)** Detection of the ΔH69/V70 (red bars) or WT sequence (blue bars) in positive SARS-CoV-2 patient samples. The positive control (patient sample with the ΔH69/V70) is displayed as green bar and the negative control as grey bar. (**G)** Prevalence of top ten SARS-CoV-2 variants in Denmark (based on pangolin lineage assignments using WGS-derived consensus genomes) from 12^th^ Jan 2021 to 8^th^ Mar 2021. **I)** Large scale screening of positive SARS-CoV-2 patient samples in the ΔH69/70 RT-qPCR in the period from 12^th^ of Jan to the 8^th^ of March 2021 (**H)** Frequency of ∆H69/70V and N501Y mutations in Denmark from 12^th^ Jan to 8^th^ Mar 2021, as determined from WGS consensus genomes obtained in this period. Mutations are relative to Wuhan-Hu-1/2019 (Genbank Accession: MN908947). **A**rrow bars in **E** and **F** indicate SEM for two technical replicates.

### Multiplexed RT-qPCR v.1

As a first step in the multiplexed RT-qPCR v.1 we developed a primer/probe pair detecting the ΔH69/V70 and WT sequence, respectively (**Fig. 2C-D)**. The limit of detection of the ΔH69/V70 RT-qPCR was 5 copies/µl for the ΔH69/V70 performing a dilution serious with a PCR standard TWIST control (Alpha B.1.1.7) (**Fig. 3E/ Suppl. Tab.2**). PCR-positive SARS-CoV-2 patient samples with paired consensus genomes from WGS were included into the ΔH69/V70 RT-qPCR. The ΔH69/V70 or WT nucleotide sequence was detected independent of the concentration of the SARS-CoV-2 sample (amount of SARS-CoV-2 RNA included per sample into the PCR) (**Fig. 2F**) and could be detected in samples of the Alpha B.1.1.7, B.1.258 and B.1.1.298 variants, where this key mutation is present (**Suppl. Fig. 1A-C)**. The ΔH69/V70 RT-qPCR correctly detected the ΔH69/V70 in SARS-CoV-2 positive samples and did not amplify samples positive for respiratory tract viruses other than SARS-CoV-2 (10/10 samples) (**Suppl. Tab. 3**). After successful validation, this RT-qPCR was incorporated as a part of the national surveillance program in Denmark driven by TestCenter Denmark, as a large-scale screen for SARS-CoV-2 variants harbouring ΔH69/V70 (starting on December 18, 2021). By mid-February 2021 the Alpha B.1.1.7 variant was the most prominent variant in Denmark (**Fig. 2G**) and at the end of March, about 80% of all SARS-CoV-2 patient samples were tested positive for the ΔH69/V70 deletion (**Fig. 2I**), which was confirmed by WGS (**Fig. 2H, Suppl. Fig. 1D**). Therefore, it was investigated if the ΔH69/V70 RT-qPCR could be multiplexed, which would then allow for the incorporation of further mutations present in other Variants of Concern. While the Alpha (B.1.1.7) variant was the most dominant variant in that time period, Beta (B.1.351) and Gamma (P.1) were still circulating in Denmark (**Suppl. Fig 1 D**). As a first step, the ΔH69/V70 RT-qPCR ran together with the diagnostic SARS-CoV-2 E-Sarbeco PCR (E-gene)^29^. The sensitivity of the ΔH69/V70 RT-qPCR was found not to be reduced when multiplexed with the E-Sarbeco RT-qPCR (**Suppl. Fig. 1E**). In conclusion, the ΔH69/V70 RT-qPCR was determined to be sensitive and specific for the detection of the ΔH69/70 as well as insensitive to multiplexing. To detect further key-mutations present in SARS-VOCs (Alpha/Beta/Gamma/Delta) and other variants of interest, we developed primers and probes to detect the L452R, E484K and N501Y mutations. Compared to the ΔH69/V70 deletion where the probe targets a stretch of a deletion of six nucleotides, the probes for the three key mutations listed above differ only by one nucleotide substitution within the *S*. Therefore, we increased their binding affinity to the mutations or the WT sequence by modifying the probes as black whole quencher plus-(BHQplus), locked nucleic acid-(LNA) or minor grove binding (MGB) conjugated probes. For the different RT-qPCRs, we tested for each mutations all primer and probe combinations, with all three probe modifications. For the N501Y mutation e.g., the MGB-conjugated probes for the N501Y mutation in the RT-qPCR were observed to be superior to locked nucleic acid (LNA) - conjugated probes, where a specific signal was detected for either the mutation or WT sequence. In contrast, the LNA probes in the N501Y RT-qPCR detected the right mutations present in the variants, but additional allelic discrimination analysis was needed to discriminate between the intensity of the signal for the mutation or the WT probe at a Ct of 45 (**Suppl. Fig. 2A-D)**.

**Figure 3.**
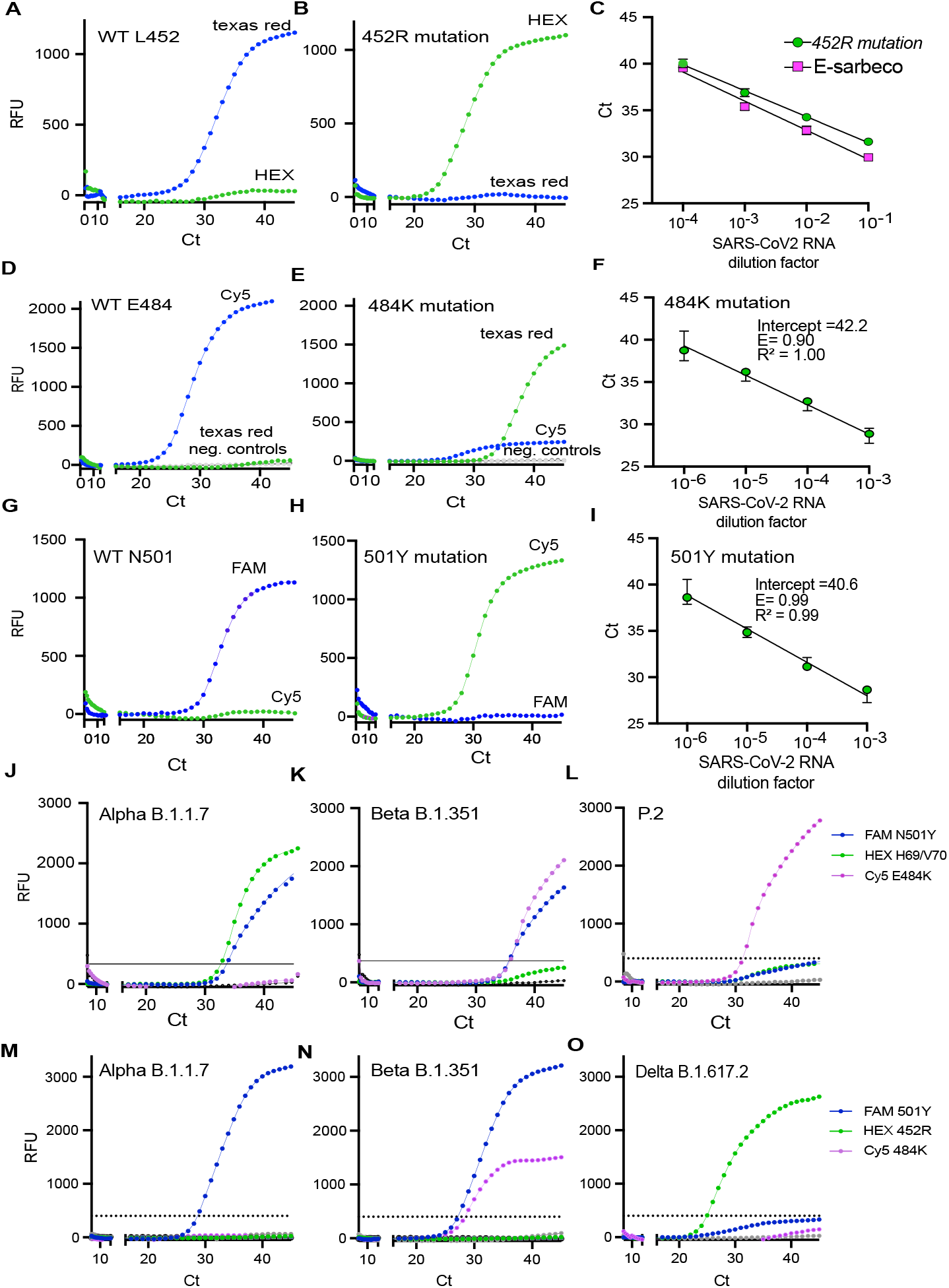
Primer and probes performance for the L452R, E484K and N501Y mutations and screening of patient samples positive for SARS-CoV-2 with different mutations of concern present in the multiplexed RT-qPCRs v.1 and v.2. **A-B)** BHQplus-conjugated probes detecting the L452 WT SARS-CoV-2 nucleotide sequence the 452R mutation. **C)** Dilution row of a patient sample with known whole genome sequence information for the delta variant (B.1.617.2) tested in parallel in the L452R- and E-sarbeco RT-qPCR. **D)** MGB-conjugated probes detecting the E484 WT SARS-CoV-2 nucleotide sequence and the 484K mutation. **F)** Dilution row of the TWIST control (Gamma P.1) included into the E484K RT-qPCR. **G)** MGB-conjugated probe detecting the N501 WT SARS-CoV-2 nucleotide sequence and the Y501 mutation. **I)** Dilution row of the TWIST control (Alpha B.1.1.7) included into the N501Y RT-qPCR. **J-L)** Detection of three key mutations ∆H69/V70, E484K and N501Y in patient samples with known whole genome sequence information identified as Alpha (B.1.1.7), Beta (B.1.351) and P.2 variants by the multiplexed RT-qPCR **v.1. M-O)** Detection of three key mutations L452R, E484K and N501Y in patient samples with known whole genome sequence information identified as Alpha (B.1.1.7), Beta (B.1.351) and Delta (B.1.617.2) variants by the multiplexed RT-qPCR **v.2**. Arrow bars in **C, F** and **I** indicate SEM for two technical replicates.

For the L452R mutation, BHQ plus conjugated probes were found to be absolutely specific compared to the LNA- and MGB conjugated probes (**Fig. 3A-B**). The limit of detection for L452R was determined by a dilution series of a patient sample with known sequence information for the delta variant (B.1.617.2) and tested in parallel in the L452R RT-qPCR and the E-Sarbeco RT-qPCR (**Fig. 3C**). The L452R RT-qPCR was about 2-fold less sensitive than the E-Sarbeco RT-qPCR (**Fig. 3C**).

The best results for the E484K mutation were gained using MGB - conjugated probes that were refined to generate a signal specific to mutation or WT nucleotide, respectively (**Fig. 3D-E, G-H**). The limit of detection for the E484K RT PCR was found to be 52 and 5 copies/µl respectively, performing a dilution series with the TWIST control Beta B.1.351 and Gamma P.1 (**Fig. 3F, I, Suppl. Tab.1**).

As a signal detected was specific either for the key mutations or the WT sequence, it was possible to only include the probes detecting the key mutations (L452R, E484K and N501Y) or the ΔH69/V70 into the multiplexed RT-qPCRs **v.1** and **v.2** (**Fig. 3J-O**). The probe for the ΔH69/70 was further modified as a Zen-conjugated probe in the multiplexed RT-qPCR v.1 to increase the signal intensity for this probe competing with the MGB-conjugating probes for E484K and N501Y mutations. Testing SARS-CoV-2 positive patient samples with known whole genome sequence information in the multiplexed RT-qPCR (**v.1**), the key mutations ΔH69/V70, E484K and N501Y present in the Alpha (B.1.17), Beta (B.1.351), B.1.5125 and P.2 were detected simultaneously if present in all patient samples (23/23) (**Fig. 3J-L**) (**Suppl. Tab 4**).

The limit of detection for the different mutations was moderately reduced to around 50 copies/µl for the different mutations in the multiplexed RT-qPCR v.1 (**Suppl. Tab.6**). To determine the specificity of the RT-qPCR v.1 we tested samples containing respiratory tract viruses other than SARS-CoV-2. Five positive signals could be detected for samples of respiratory tract viruses, but with a CT higher than 38 in the multiplexed **RT-qPCR v.1** (**Suppl. Tab. 5**). Repeating the experiments twice with the same samples in **RT-qPCR v.1** resulted in a negative result (**Suppl. Tab.5**). As the limit of detection for the multiplex **RT-qPCR v.1** was at a CT of 37 for the N501Y mutation, positive Ct values > 38 should be considered as negative (**Suppl. Tab. 6**).

### Multiplexed RT-qPCR v.2

As proof of concept and to investigate the robustness of the multiplexed RT-PCR we investigated if the ΔH69/V70 could be replaced by the L452R mutation, present in the delta variant (B.1.617.2) in the multiplexed **RT-qPCR v.2**. As it is recommended to limit the number of MGB-conjugated probes in a multiplex RT-qPCR, we combined the two MGB-conjugated probes for the E484K and N501Y mutations with a BHQ-plus-conjugated probe for the L452R mutation. With this approach the three key mutations L452R, E484K and N501Y could be simultaneously detected in 31/31 samples with known sequence information for the alpha (B.1.1.7), beta (B.1.351) and zeta (P.2) variants in the multiplexed **RT-qPCR v.2** (**Suppl. Tab.3**).

The multiplexed small-scale RT-qPCR platform offers a flexible and fast detection system to rapidly identify key mutations present in SARS-CoV-2 VOCs and mutations of interest. Notably, new key mutations can be accommodated by exchanging one of the existing sets. This forms the basis of a flexible detection platform where three key mutations can be detected in parallel in the multiplexed RT-qPCRs for small-scale screening.

### LARGE SCALE SCREENING OF THE VARIANT RT-qPCR AS PART OF THE NATIONAL SURVEILLANCE PROGRAM IN DENMARK

The same primer and probes designed for the four key mutations (ΔH69/V70, L452R, E484K and N501Y) included into the multiplexed RT-qPCR for small-scale screening were further validated for large-scale screening, implemented to support the national surveillance program in Denmark in addition to WGS, supporting the public health system to delay the emergence of VOC. Large-scale screening consisted of RT-qPCRs running in parallel on a 384-well plate allowing for parallel detection of the four key mutations. The two key mutations, ΔH69/V70 and N501Y run as multiplexed RT-qPCR in large scale, were detected in 17/17 (100 %) of patient samples with known sequence for the Alpha (B.1.1.7) and Beta (B.1.351) variants (**Suppl. Tab. 7**). The L452R and E484K mutations were correctly detected in single RT-qPCR reactions in 18/18 (100%) and 31/31 (100 %) patient samples respectively, with known sequence information for the Alpha (B.1.1.7), Beta (B.1.351), Delta (B.1.617.2), Zeta (P.2) or B.1.525 variants (**Suppl. Tab. 8-9**). The ΔH69/V70/N501Y, L452R and E484K RT-qPCRs for large-scale screening were specific, as these did not yield a positive signal in samples positive for common respiratory tract viruses other than SARS-CoV-2 (**Suppl. Tab.2**). Based on these results, the RT-qPCRs were implemented into the large-scale screening at TestCenter Denmark, where the sensitivity and specificity were tested in comparison to WGS data (**Fig. 4A-C**).

**Figure 4.**
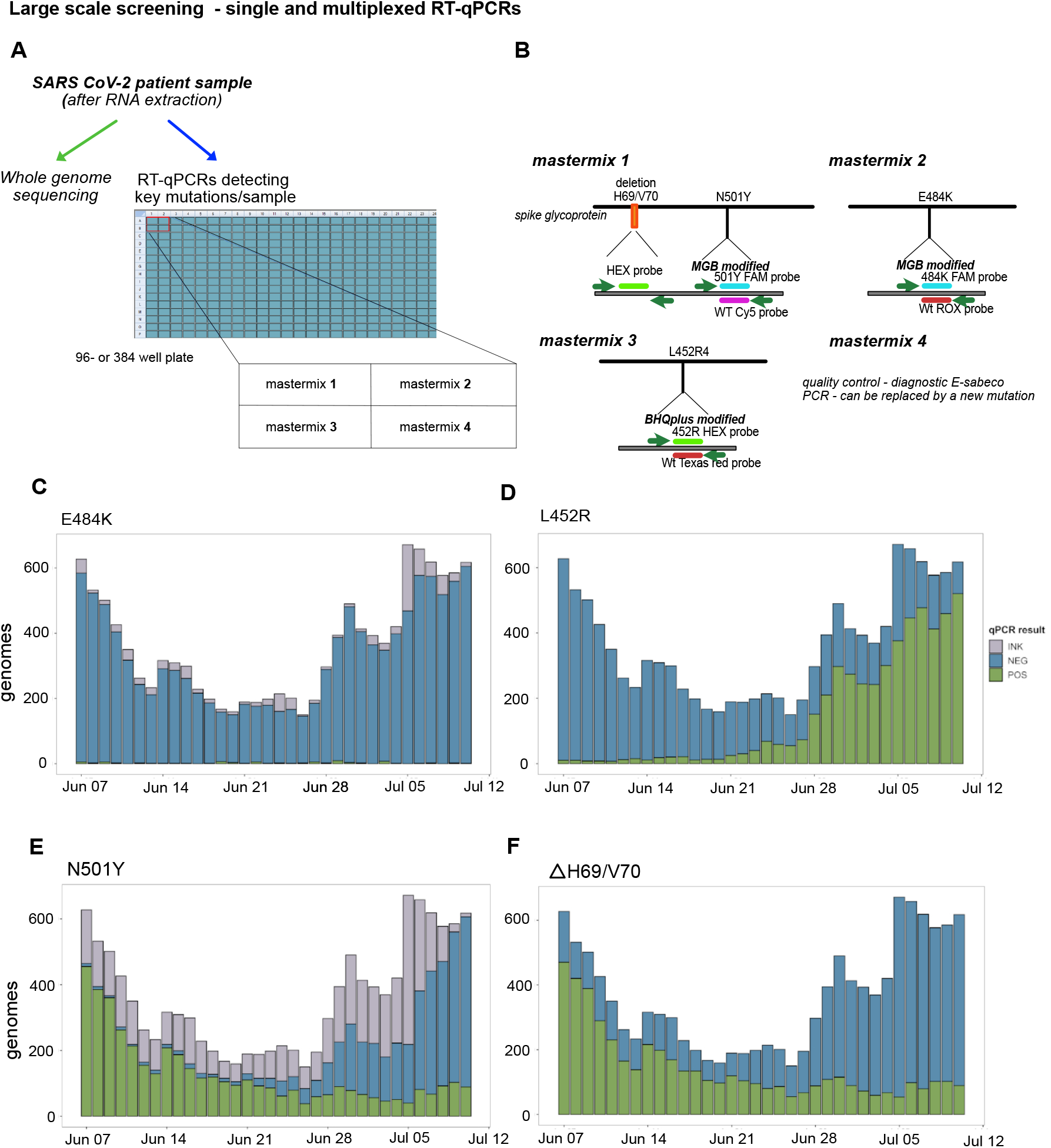
Large scale screening of four key mutations. **A)** Schematic overview of large-scale screening. **B)** Primer and probes included into the multiplexed and single PCR running in the 384-well plate format. **C)** RT-qPCR results from large-scale screening for each target mutation: E484K (upper left), L452R (upper right), N501Y (lower left), ∆H69/70V (lower right) shown as positive (POS, green), negative (NEG, blue) or inconclusive (INK, grey).

To validate the RT-qPCR implemented as large-scale screening, results from 9572 positive samples were tested both in the RT-qPCR and by WGS over a five-week period from the 7^th^ of June 2021 to the 11^th^ of July 2021 were compared. This period was selected due to the presence of all four key mutations of interest in genomes sequenced as part of this national surveillance strategy. It is also during this period the dominant B.1.1.7 (Alpha) SARS-CoV-2 variant^30^ was seen to be replaced by the more transmissible^31^ B.1.617.2 (Delta) variant in Denmark. This then allowed for a rigorous test of the RT-qPCR strategy due to the presence and absence of these key mutations amongst these multiple variants (**Fig. 1A-B**). A daily range of 150 to 671 samples were analysed by multiplex RT-qPCR during this period and results were characterised as either positive (POS) or negative (NEG) for a given key mutation (**Fig. 4C**). It was observed that there was a small number of inconclusive results amongst the E484K RT-qPCRs, as well as a more noticeable number of inconclusive results in the N501Y RT-qPCR which could be attributed to probe manufacturing issues beyond our control – replacement of the probes resulted in a significant reduction in the number of inconclusive results from this reaction (**Fig. 4C/**lower left panel), Jul 10^th^ to Jul 11^th^, 2021). This probe was found to be more sensitive to the concentration of the samples, thus samples with high CT values in the initial E-Sarbeco based analysis had a tendency to yield inconclusive results. Thus, the N501Y RT-qPCR was more sensitive to minor variations in batch quality. In order to validate all RT-qPCR results and determine the specificity and sensitivity of these primer/probe combinations, WGS consensus genomes from the same samples were used as a reference standard.

WGS was performed on all positive samples during the study period using the ARCTIC Network’s PCR scheme v3 (see Materials and Methods) and the aligned *S* gene sequences from the resulting consensus genomes were used to validate the results of each of the three RT-qPCR reactions, by comparison of translated codons to RT-qPCR results at each position encoding the four key mutations of interest in this study. Validation was performed on samples where both a valid RT-qPCR result and a consensus genome sequence was obtained, a number which differed for each of the four key mutations for various technical reasons anticipated at this scale (see above). The validation results (**Fig. 5**) showed good agreement between amino acids translated from WGS and RT-qPCR results for E484K, N501Y and L452R (**Fig. 5A-C**). The determination of concordance proved less straightforward for ∆H69/V70 due to the alignment of reads around the deletion prior to consensus generation, resulting in a significant discordant fraction between the deletion and negative RT-qPCR results (**Fig. 5D**). It was also observed amongst the consensus genomes used in this validation that amino acid 452 in the spike protein was more mutable than the other positions which form this set of key mutations, with L, R, M and Q observed at this position depending on the lineage (Q484 was not observed in genomes during the selected period but has been recorded in global surveillance data).

**Figure 5.**
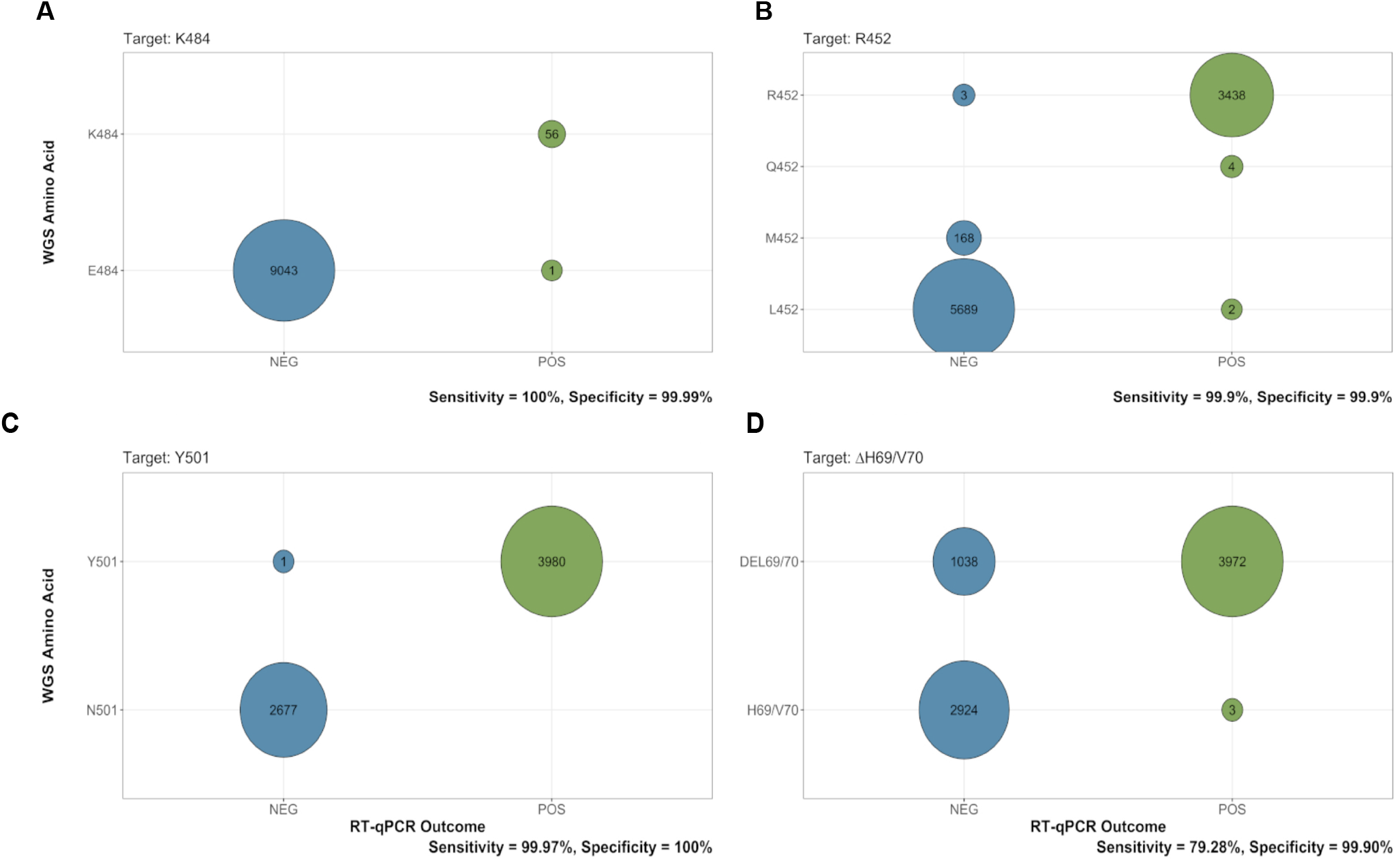
Validation of RT-qPCR results from large-scale screening. Concordance between RT-qPCR results and WGS results represented as a graphical matrix with each cell represented as a circle showing the number of samples which correspond to a positive (POS, green) or negative (NEG, blue) RT-qPCR result (horizontal axis) and a given amino acid derived from WGS consensus genomes (vertical axis) for E484K (upper left panel), L452R (upper right panel), N501Y (lower left panel), ∆H69/70V (lower right panel). Sensitivity and specificity for each RT-qPCR shown at bottom right of each panel.

In order to meaningfully compare and describe the relative performance of the RT-qPCR strategy from the results of the large-scale screen, as well as to determine the true-positive or true-negative rate of these combinations of primers and probes, the specificity and sensitivity of each primer/probe combination was determined using established methods used to characterise diagnostic testing^32^. Using the validation of the RT-qPCR with WGS as a reference standard, specificities and sensitivities were calculated for all primers and probes for the four key mutations, and it was observed that all four RT-qPCRs were highly specific (>99.9%), and three out of four assays were highly sensitive (>99.9%) (**Fig. 5 A-D**). The sensitivity of the probe for detection of ∆H69/V70 was observed to be reduced (79.28%) due to a significant number of deletions in WGS, which were assayed to be negative by RT-qPCR; however, given the challenge of read alignments around genomic regions containing insertions or deletions, this was postulated to be largely due to the determination of the deletion in WGS consensus genomes. In addition to the specificity and sensitivity of the primer/probe combinations, the Positive Predictive Value (PPV) and Negative Predictive Value (NPV) of these combinations was also determined, which indicates the ability of a diagnostic assay or test to accurately detect a condition or in this case, mutation^32^. The determination of PPV and NPV takes into account the specificity and sensitivity of the primer/probe combinations as well as the prevalence of the four key mutations amongst the sequenced SARS-CoV-2 genomes during the study period:

It was determined that all four primer/probe combinations had a PPV of at least 97 %, and a NPV of 99.9 % for three out of four of these, with the ∆H69/V70 assay having a NPV of 75.43 % (**Fig. 5 A-D**). From the results of the large-scale screening, it can be seen therefore that the specificity and sensitivity as well as the PPV and NPV all point towards the viability of this RT-qPCR strategy in a large-scale diagnostic setting.

In summary, we developed a RT-qPCR system for large-scale screening of four key mutations in parallel that is highly specific and sensitive, validated by a comparison of the qPCR and WGS data from 9572 samples that were tested in parallel.

## DISCUSSION

RT-qPCRs platforms for small and large-scale screening can support the detection of mutations of concern present in SARS-CoV-2 variants. This is of special interest for countries lacking an infrastructure for large-scale WGS sequencing, the golden standard for SARS-CoV-2 variant surveillance. Here, a detection system is needed that is fast, robust and flexible and, which enables the detection of known diagnostic mutations almost in real-time after sample collection, as we showed in this study.

Here, we describe validated and advanced RT-qPCR platforms for small and large-scale screening that can simultaneously detect mutations of concern within the *S* of SARS-CoV-2, with a fast turnaround time for large-scale screening of 12-24h to report to the public health system. In comparison to commercially available systems to detect mutations of concern, the RT-qPCR platforms can be established fast and new mutations can be implemented; an important advantage to follow the course of a pandemic. It is a transparent system, where troubleshooting is possible without depending on the knowledge from a company and it can be adjusted to the existing infrastructure of the laboratory for large-scale screening and data evaluation.

Moreover, the RT-qPCR platforms are at low cost of about 10 DKK (2 USD) per reaction, and can therefore be establish in countries without the resources for WGS in large-scale RT-qPCR is a fast, standard method for SARS-CoV-2 detection and has been established at the start of the pandemic in January 2020^29^. The standardised protocol for SARS-CoV-2 RT-qPCRs makes it easy to implement into diagnostic laboratories worldwide, where the equipment needed is commonly present. This could be of advantage compared to new methods such as RT-LAMP or CRISPR, which have been described for SARS-CoV-2 detection, delivering faster test results and can be applied without extensive laboratory equipment as RT-qPCRs^33–35^.

Currently there are only limited studies on RT-LAMP for commercial point of care^3^. Moreover, CRISPR is still in its infancy^36^ and has been shown to be less sensitive compared to RT-qPCR^3^. As most diagnostic facilities worldwide do not possess access and knowledge to establish these technologies, opposing to RT-qPCR that is a universal standard method, RT-qPCRs are still the method of choice for most diagnostic laboratories. Based on recent advances in modifications of conjugated-probes, RT-qPCRs can be designed to detect mutations within the SARS-CoV-2 genome consisting out of a single nucleotide polymorphism (SNP).

For small-scale screening the multiplexed RT-qPCR was developed using a Luna Probe One-Step RT-qPCR Mix, which offers the possibility to increase the input template concentration, for amplification targets with a low RNA concentration, as it is four times more concentrated. However, this was not an advantage when establishing the multiplexed RT-qPCRs **v.1** and **v.2**, as SARS-CoV-2 RNA concentrations vary among patient samples and can be too high from start leading to artificial signals. In contrast, adjusting the primer and probe concentrations for each mutation resulted in a highly specific and sensitive detection of the corresponding mutation present in the different SARS-CoV-2 variants (**Fig. 3J-O, Suppl. Tab. 4**). Moreover, by reducing the number of probes in the multiplexed RT-qPCR we could maintain a sensitive system for diagnostic use, by including one probes for each mutation. This was possible by designing and testing combinations of primer and MGB-, LNA- or BHQplus-conjugated probes that yield a specific signal for the mutation and WT sequences, respectively. The best performance for each primer/probe pairs is empirical and should be tested for all possible probe modifications (LNA, MGB and BHQplus), as the result is dependent on the nucleotide sequence of the mutation or WT sequence. So far, only mutations of concern within the *S* were included into the multiplexed RT-qPCR platform for small-scale screening but running the ΔH69/70 RT-qPCR as multiplexed PCR together with the clinical E-sarbeco RT-qPCR did not reduce the sensitivity of the PCR (**Fig. 2E**).

As up to five targets can be included into the multiplex RT-qPCR using the Luna Probe One-Step RT-qPCR Mix, additional targets located in other loci of the SARS-CoV2 genome than *S* could be included. We did not test the maximum number of targets that could be included into RT-qPCR platform, as this was out of the scope of this study, but this could be of interest for future studies.

Large-scale RT-qPCR screening of mutations present in VOCs required that a number of technical and analytical considerations can be fulfilled: 1) the RT-qPCR must be highly specific and sensitive to minimise or avoid false positives, 2) it should be of sufficient robustness to allow for massive scalability required in a pandemic, 3) it must not interfere with the diagnostic PCR to detect SARS-CoV-2 to reduce the risk of potential PCR contamination 4) it requires liquid handlers in order to be viable from a practicable standpoint and 5) an advanced, automated evaluation system is needed to detect the erroneous results. Here we describe a large-scale RT-qPCR platform that meets the technical and analytical considerations outlined above. The current design is based on sample preparation in a 96-well format and subsequent RT-qPCR in 384-format. This allows each sample to be analysed by four separate sets of primers and probes, which enables the analysis of four mutations for up to 92 samples and four controls (one negative and three positive) in parallel in a single run. The system is flexible as the combination of target mutations can be adjusted over time in accordance with current needs. The handling of data calls for automated data processing and variant calling which is due the large amount of data in each run and the complex calling algorithms. Inconclusive results can pose a challenge with regards to variant calling. When one or more of the mutations are inconclusive, it is not always possible to make an unequivocal variant call. In our set-up we have opted to report the detected mutations. In these cases, prominent mutations with putative biological functions in various VOCs were reported, rather than variants of concern and interest.

From the large scale-screen it was determined that the RT-qPCR platform described in this study is generally of very high specificity and sensitivity and performs well in terms of its PPV and NPV, indicating its utility in such large-scale diagnostic screens. The period for the large-scale screening and validation was specifically chosen to interrogate the robustness of this system in a pandemic transition period with ongoing lineage replacement; such a period involves the waning of certain variants such as the Alpha (B.1.1.7) and its signature mutations ΔH69/V70 and N501Y, along with the rise of a different variant like Delta (B.1.617.2) with a different signature mutation (L452R). In order to have diagnostic value, surveillance mechanisms which track these exclusive signatures, and which do not yield full genomes, must have adequate sensitivity and specificity to be able to adequately distinguish between such signature mutations (this is also aided by the E-Sarbeco PCR result, being the primary diagnostic method used to determine a SARS-CoV-2 infection). In that respect, the sensitivity and specificity of this system is excellent, only falling short in sensitivity in one assay (∆H69/V70) due to distinct technical issues, all of which revolve around the WGS reference standard and not the RT-qPCR itself. Firstly, the challenge of read alignment around genome deletions leads to ambiguous base-calls around these regions. Secondly, in large scale amplicon-based genomic surveillance, dropouts are not a rare occurrence, and a certain degree of N-counts is therefore considered permissible (typically less than 5-10 % of the consensus genome). Tracts of Ns around this region were observed around the deletion and this was largely responsible for the challenges of identifying a deletion from WGS consensus genomes. However, this was not the case for single SNPs leading to non-synonymous substitutions as seen with N501Y, E484K and L452R. Interesting insights into the specificity and the sensitivity of the RT-qPCR system were also observed in the results around the L452R mutation, given that position L452 in the spike protein exhibited more than a single amino acid change during the pandemic and indeed the timeframe of the large-scale screening performed. The validation showed that all samples with L452Q in the spike protein recorded a positive result from the RT-qPCR whereas L452M exclusively recorded negative RT-qPCR results. Given that the codon observed from WGS encoding for Q was *cag* and the corresponding codon encoding for L at the same position was *cgg*, this could be considered unsurprising, also given that Q452 was not an anticipated mutation and therefore was not considered in the design of the probes. Given that the codon *atg*, which is more distant from *cgg* and which encodes for M at this position, was not detected by the L452R-specific probe, this alludes to the specificity and sensitivity of the RT-qPCR probe at position 452.

One of the major arms of pandemic control seen in this pandemic revolves around the screening and isolation of SARS-CoV-2 infected individuals in order to limit community spread of infections. The screening and isolation of individuals is therefore time sensitive and requires a rapid turnaround, especially where the control of variants or mutations of concern are a priority. While WGS of positive samples affords the accurate identification of these variants or mutations in order to enable their tracking and therefore control, this entails a longer turnaround time and greater cost in terms of reagents, equipment and expertise. The use of RT-qPCR systems such as the one described in this study allows for rapid identification of mutations of concern, which in turn enables near-real-time tracking of these and correspondingly, rapid decision-making around testing, contact tracing and isolation. This enabled the rapid reaction of the public health system in Denmark to the detection of VOCs, with the added benefit of gaining time to implement its vaccination schedule; being in line with modelling showing that minimising testing delay, had the largest impact on reducing onward transmissions^37^. The flexibility of this system also allows for multiplexing to detect multiple mutations and the incorporation of new primers and probes in response to the dynamics of the SARS-CoV-2 pandemic. In addition, the specificity and sensitivity of this system show that it is robust and therefore suited to diagnostic requirements in a pandemic. Taken together, these characteristics make this RT-qPCR system an ideal candidate for laboratories looking to detect mutations of concern in the SARS-CoV-2 pandemic. The current shift in our consideration of the pandemic (towards endemicity) suggests that such monitoring and screening might have to last a considerably longer time, making this system extremely viable in the long-term, and indeed in future outbreaks and pandemics.

## Material and Methods

### Ethics

Exemption for review by the ethical committee system and informed consent was given by the Committee on Biomedical Research Ethics - Capital region in accordance with Danish law on assay development projects.

### Virus isolation

SARS-CoV-2 viral isolates representative of VOC (Delta variant B.1.617.2, Alpha variant B.1.1.7 and Beta variant B.1.351 were isolated from PCR-positive throat swabs collected in phosphate buffered saline (PBS) from community testing facilities (Test Center Denmark) and BioBank Denmark, which form part of the Danish national surveillance program^4^. The primary isolation was performed in 24-well culture plates with 5×10^4^ Vero E6 cells/well seeded the day before. Cells were washed once with PBS, and 150-250 µL of swab material and 150-250 µL infection media [Dulbecco’s Modified Eagle Medium (DMEM) with 1% Penicillin/Streptomycin] were added to each well. After 1h incubation at 37°C/5% CO_2_, 1 mL/well of propagation media [DMEM with 1% Penicillin/Streptomycin, 5% foetal calf serum] was added, and the cultures were further incubated until cytopathic effect (CPE) was observed. Isolations performed later during the pandemic used additional 1.5 µg/mL Amphotericin B in the propagation media. All cell culture reagents were obtained from Gibco, ThermoFisher Scientific, Waltham, MA, USA. Upon CPE, supernatants were aliquoted and frozen at -80°C. Subsequent passages to expand virus stocks were performed in 75 cm^2^ flasks seeded with 1.5×10^6^ Vero E6 the day before. 25 µL of primary isolate supernatant was used as inoculum in the presence of 2 mL infection media. After 1h at 37°C/5% CO_2_ incubation, flasks were supplemented with 10 mL of propagation media (without Amphotericin) and incubated until CPE was obtained.

Supernatants were then clarified by centrifugation for 5 min at 300 x g and stored as single use aliquots at -80°C.

### RT-qPCR validation standards and patient samples

For determining specificity and sensitivity of the SARS-CoV-2 Variant PCR assays, the following materials were used:

Diagnostic samples positive for the common respiratory pathogens Human Coronavirus 229E, HKU1, NL63 and OC43, Adenovirus and Rhinovirus, was obtained as extracted nucleic acids from the human diagnostic Virus PCR laboratory at Statens Serum Institute, Denmark, and were all previously confirmed by PCR to be positive at high concentration (Ct <<30) for respective pathogens.

Extracted Influenza virus RNA from viruses cultured in Madin Darby Canine Kidney (MDCK) cells (A/Christchurch/16/2010(H1N1), pdm09-like virus, B/Phuket/3073/2013-like virus, B/Brisbane/60/2008-like virus, were all previously confirmed by PCR to be positive at high concentration (Ct <<30) for respective pathogens. The influenza reference viruses was provided by the WHO Collaborating Centre for Reference and Research on Influenza, The Francis Crick Institute, London, United Kingdom. Positive RNA controls for SARS-CoV-2 variants were obtained from extracted virus cultures and were diluted in DNase/RNase free water to generate CT values between 25-30 in the subsequent RT-qPCR.

TWIST Synthetic SARS-CoV-2 RNA controls (MT007544.1/Australia/VIC01/2020), (MT103907 England/205041766/2020), (MT104043 South African/KRISP-EC-K005299/2020) and (MT104044 Japan (IC-0564/2021) were bought from TWIST bioscience and used as PCR standards WT, Alpha (B.1.17), Beta (B.1.351), Gamma (P.1), respectively.

Selected SARS-CoV-2 VOC positive patient samples were obtained from the Danish National Biobank.

### Positive and Negative controls for the large-scale RT-qPCR platform

Positive and negative controls for the large-scale platform were run in parallel with selected patient samples throughout extraction and RT-qPCR. DPBS 1x pH 7.2 (Gibco) was used as negative control. Heat inactivated (56 °C for 45 min.) virus cultures, were used as positive control. Three Danish virus isolates were used to cover the four key mutations present in the Delta variant B.1.617.2, Alpha variant B.1.1.7 and Beta variant B.1.351 (SSI-H18).

### Nucleic acid extraction

For small scale SARS-CoV-2 patient sample screening, total nucleic acid was extracted using a MagNApure96 extraction robot (Roche) with the MagNA Pure 96 DNA and Viral NA Small Volume kit and the Viral NA Plasma SV protocol (200 µL input and 100 µL elution volume).

For positive controls, 120 µL of supernatant from SARS-CoV-2 infected cells were mixed with 120 µL of MagNA Pure lysis buffer (Roche) and extracted as small-scale SARS-CoV-2 patient samples. Positive control RNA was stored at -80°C until use. For large-scale SARS-CoV-2 patient sample screening, RNA was extracted using a Beckman Coulter Biomek i7 robot using the Beckman Coulter RNAdvance Whole blood kit (200 µL input and 50 µL elution volume).

### Primer and probe design

SARS-CoV-2 variant sequences were retrieved from positive samples identified through the national surveillance program in Denmark. Sequences were aligned and primer and probes were designed using Geneious Prime 2021.0.

Two probes were designed for each key mutation: one detecting the wildtype (WT) nucleotide sequence, and one detecting the mutation. The probe design was refined to detect the key mutations (L452R, E484K, N501Y, Δ69/V70 deletion) with only one probe in the multiplex RT-qPCRs. To ensure stable allelic discrimination analysis, probes detecting the mutations with only one nucleotide exchange were either MGB, LNA or BHQplus modified, which increases the melting temperature (Tm). The calculation of MGB probe Tm was adapted from^38^.

The primers and probes listed in Tab. 2 were synthesized by Biosearch Technologies, Denmark, except for the MGB-probes that were synthesized by Eurogentec, Belgium, and the Zen-probe, that was synthesized by Integrated DNA Technologies, Belgium. All oligos were HPLC-purified.

**Tab.2:**
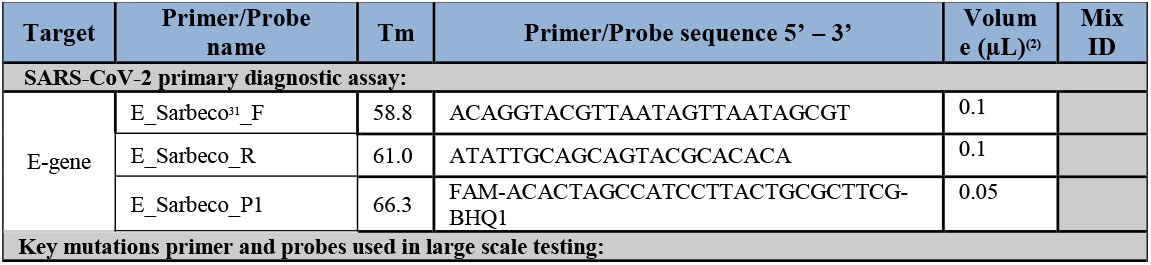
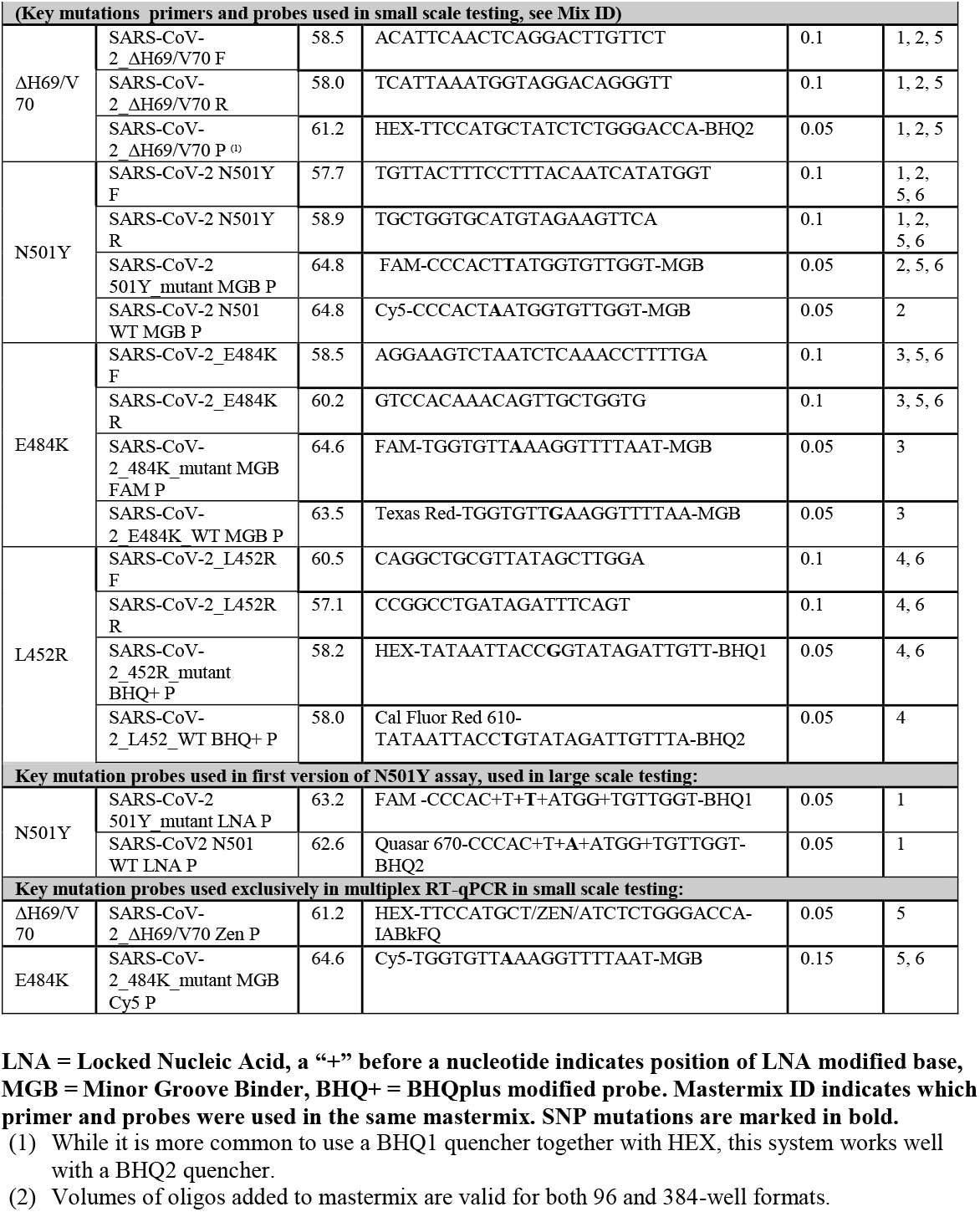
Primer and probe sequences.

**Tab 3.** Positive and Negative Predictive Values for all 4 RT-qPCR assays during period of large-scale screen (7^th^ June 2021 to 11^th^ July 2021).

| Mutation | Prevalence (%)<br>(No. of samples) | PPV (%) | NPV (%) |
| --- | --- | --- | --- |
| ΔH69/V70 | 61.1 (5010) | 99.9 | 75.4 |
| N501Y | 52.3 (5846) | 100.0 | 99.9 |
| E484K | 0.6 (60) | 97.7 | 100.0 |
| L452R | 35.9 (3441) | 99.8 | 99.9 |

Mutation prevalence estimates were calculated based on consensus sequences from 9572 positive samples (as determined by E-Sarbeco PCR) obtained from this period.

### Mastermix set-up

The primers and probes were combined in different master-mixes.

In master-mix 1-4 (large-scale screening): the mutations were detected using both the mutant probe and the wildtype probe for allelic discrimination analysis.

In master-mix 5 and 6 (small-scale screening), only probes targeting the mutations were used, and therefore no allelic discrimination analysis was needed.

#### 96-well format PCR conditions used in development phase and small-scale testing

All PCR assays were developed on a Bio-Rad CFX 96 PCR real-time PCR system. **Master-mix 1 - 4** contained 12.5 µL Luna^®^ Universal Probe One-step RT-qPCR Kit reaction buffer (NEB), 1.25 µL Luna^®^ WarmStart RT Enzyme mix, primers and probes (100 µM, volumes in table 1), DNAse/RNAse free water and 5 µL template to a total volume of 25 µL. Cycling conditions: Reverse transcription at 55 °C for 10 min., initial denaturation at 95 °C for 3 min., followed by 45 cycles of denaturation and annealing/extension at 95 °C for 15 sec. and 58 °C at 30 sec. respectively.

**Master-mix 5 - 6** contained 5 µL Luna Probe One-Step RT-qPCR 4X Mix with UDG (New England Biolabs Inc (NEB)), primers and probes (100 µM, volumes in Tab. 1), DNAse/RNAse free water and 5µL template to a total volume of 25 µL. Cycling conditions: Initial step at 25°C for 30 sec, reverse transcription at 55 °C for 10 min., initial denaturation at 95 °C for 1 min., followed by 45 cycles of denaturation and annealing/extension at 95°C for 10 sec and 58°C at 60 sec respectively.

#### Data analysis for the multiplexed RT-qPCRs used in small-scale testing

The multiplexed RT-qPCRs contain probes only targeting the mutations, ΔH69/V70, 501Y, 484K for **master-mix 5**, and 501Y, 484K, 452R for **master-mix 6**. Cut-off values were used in the multiplexed RT-qPCRs to secure the detection of only the mutation and not the WT sequence as there was no WT probe in the mix. A sample was considered positive with these criteria: Ct <38 and RFU (Relative Fluorescence Units) > 500 at Ct = 45.

#### 384-well format PCR conditions for large-scale testing

In large scale testing, the assays run on a Bio-Rad CFX 384 PCR real-time PCR system. The master-mix contained 7.5 µL Luna^®^ Universal Probe One-step RT-qPCR Kit reaction buffer (New England Biolabs Inc), 0.75 µL Luna^®^ WarmStart RT Enzyme mix, primers and probes (100 µM, volumes in table 1), DNAse/RNAse free water and 5µL template to a total volume of 15 µL. Cycling conditions were the same as for the 96-well format. Each patient sample was analysed in four PCR wells, in four parallel reactions, using master-mix 1 or 2 for detecting ΔH69/V70 and N501Y, master-mix 3 for detecting E484K, master-mix 4 for detecting L452R and in the final well the E-Sarbeco assay was used for detection of SARS-CoV-2 wildtype (E-gene). The 4 master-mixes were placed in a quadratic pattern, thus allowing easy transfer from a 96-well plate to a 384-well plate (e.g. A1 in a template plate was pipetted to A1, B1, A2 and B2 of the master mix plate). Master-mix 5 and 6 were not tested in the 384-well format.

#### Data analysis using allelic discrimination analysis in large-scale testing

PCR curves were evaluated in the Bio-RAD CFX software and Ct values and end RFU were exported in csv files. The files were imported into the laboratory database where all data analysis was performed. For the ΔH69/V70 deletion, detection was based on Ct values (deletion detected is Ct = 12-38). For the the mutations N501Y, E484K and L452R, detection was based on allelic discrimination where the end RFU values were utilized to determine the presence of a mutation (see Suppl. Tab. 1). A sample was considered positive with these criteria: Ct <38 and RFU > 200 at Ct = 45. The RFU cut-off value was used in the 384-well PCR-format as a quality control step, in case one of the probes in the allelic discrimination pair failed.

#### Whole genome sequencing

Whole genome sequences were generated by The Danish COVID-19 Genome Consortium (DCGC) from PCR-positive samples collected between 6^th^ June and 11^th^ July 2021. Samples were selected using Ct cut off values between 30 – 38^30^. The bulk of the samples were sequenced using the ARTIC Network tiled PCR scheme V3 via the COVIDseq Assay [Illumina], Artic Network nCoV-2019 sequencing protocol v2 *(dx*.*doi*.*org/10*.*17504/protocols*.*io*.*bdp7i5rn* [Oxford Nanopore], or a custom DCGC protocol (*dx*.*doi*.*org/10*.*17504/protocols*.*io*.*bfc3jiyn*)[Oxford Nanopore], adapted from the Artic Network protocol. Data pre-processing and consensus genome generation was performed using Illumina-specific (github.com/connor-lab/ncov2019-artic-nf, v. 1.3.0) or Oxford Nanopore-specific (github.com/artic-network/fieldbioinformatics, v. 1.2.1) consensus pipelines. Consensus genome mutation calling with reference to Wuhan-Hu-1/2019 (Genbank Accession: MN908947) was performed with Nextclade CLI (github.com/nextstrain/nextclade, v. 1.2.0) and lineage designations were performed using pangolin (github.com/cov-lineages/pangolin, v. 3.1.3) with the accompanying pangoLEARN model (github.com/cov-lineages/pangoLEARN, v. 1.2.6).

#### RT-qPCR Validation

Nucleotide sequences corresponding to the Sof consensus genomes derived from WGS were aligned using the MAFFT version 7.480 (*mafft*.*cbrc*.*jp*), utilizing the FFT-NS-2 algorithm with a maximum of 1000 iterations^40,41^. Alignments were viewed and processed in Jalview 2.11.1.4 (*jalview*.*org*,^42^) and codons encoding key mutations were extracted, translated and compared to RT-qPCR results. From here, sensitivity, specificity, Positive Predictive Values (PPV) and Negative Predictive Values (NPV) were calculated for each set of primers and probes used in RT-qPCR assays. Positive and Negative Predictive Values were calculated according to the following formulas:

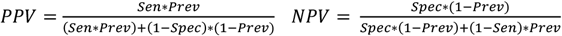

where *Sen* = sensitivity, *Spec* = specificity and *Prev* = prevalence calculated from WGS consensus genomes. All analyses were performed in Rstudio version 1.4.1717 using R version 4.1.1 and using the packages *tidyverse* (1.3.1), *seqinr* (4.2-8), *lubridate* (1.7.10), *ggplot2* (3.3.4), *cowplot* (1.1.1), *zoo* (1.8-9) and *ggpubr* (0.4.0).

#### DATA analysis

We used standard curves to determine the SARS-CoV-2 detection threshold for each assay and to calculate the viral load in each sample. We used the SARS-CoV-2 variant specific TWIST control with a known concentration (copies/µl) and diluted 1:10 in a seven-step dilution series. The median Ct-values and the interquartile ranges were calculated based on biological duplicates with technical duplicates. The threshold was based on the intercept of the linear regression line of the standard dilutions. Furthermore, the number of virus particles were estimated based on the logarithmic regression function of each assay’s standard dilution series.

## Data Availability

All data produced in the present study are available upon reasonable request to the authors.

## Figure legends

**Supplementary Figure 1.**
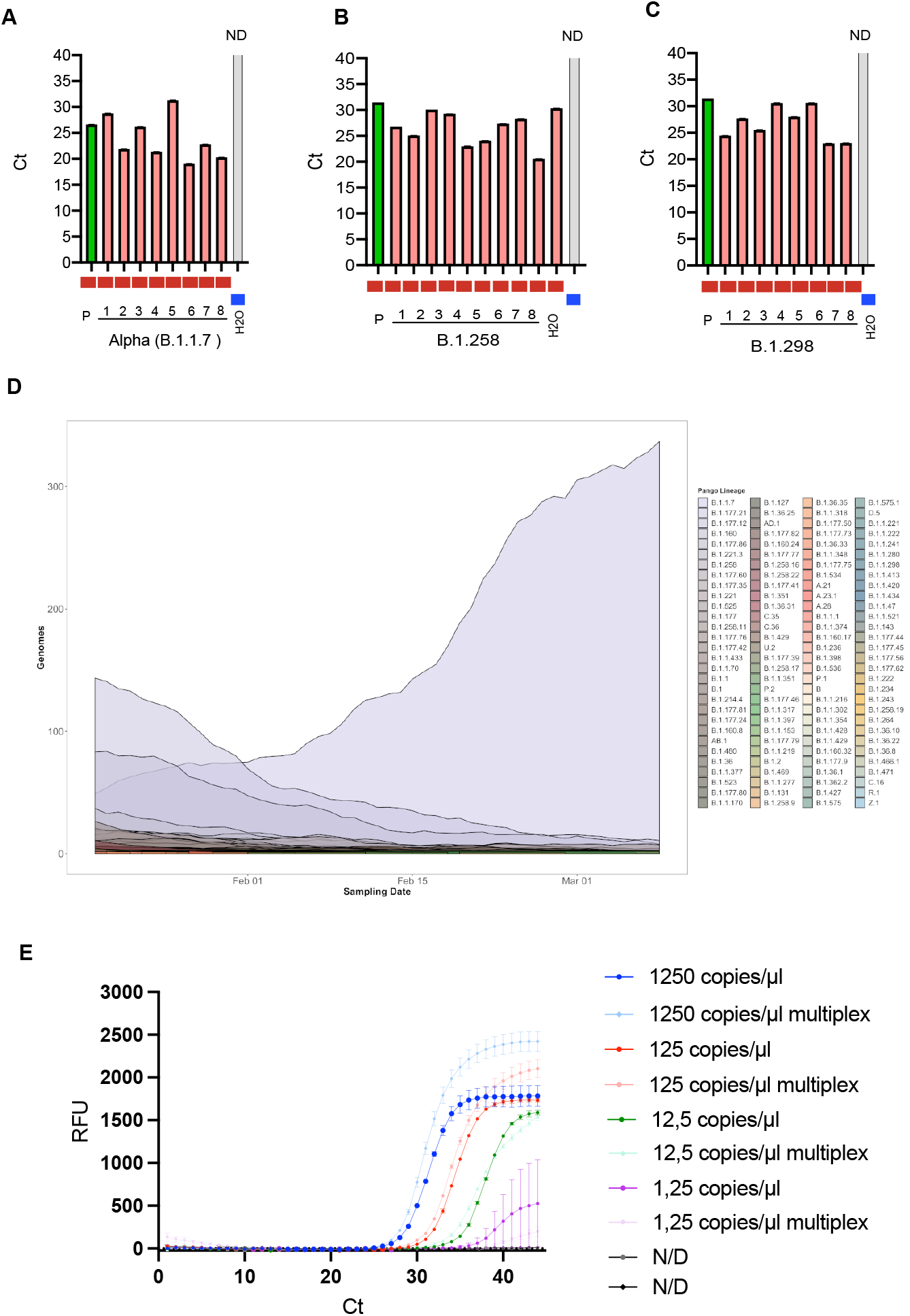
Detection of the H69/70V deletion by RT-qPCR. **A-C)** Detection of the ΔH69/V70 in patient sample with known whole genome sequence information identified as Alpha (B.1.1.7), B.1.258 and B.1.298 variants (red bars). Positive control (green bar) of sample with known sequence information positive for the ΔH69/70 and the negative control (grey bar). **D)** Prevalence of all SARS-CoV-2 variants in Denmark (based on pangolin lineage assignments using WGS-derived consensus genomes) from 12^th^ Jan 2021 to 8^th^ Mar 2021. **E)** Dilution of the TWIST control (WT SARS-CoV-2) and detection of the H69/V70 WT sequence by the H69/V70 RT-qPCR or the multiplexed H68/V70_E-sarbeco RT-qPCR. Arrow bars in **E** indicate SEM for two technical replicates.

**Supplementary Figure 2.**
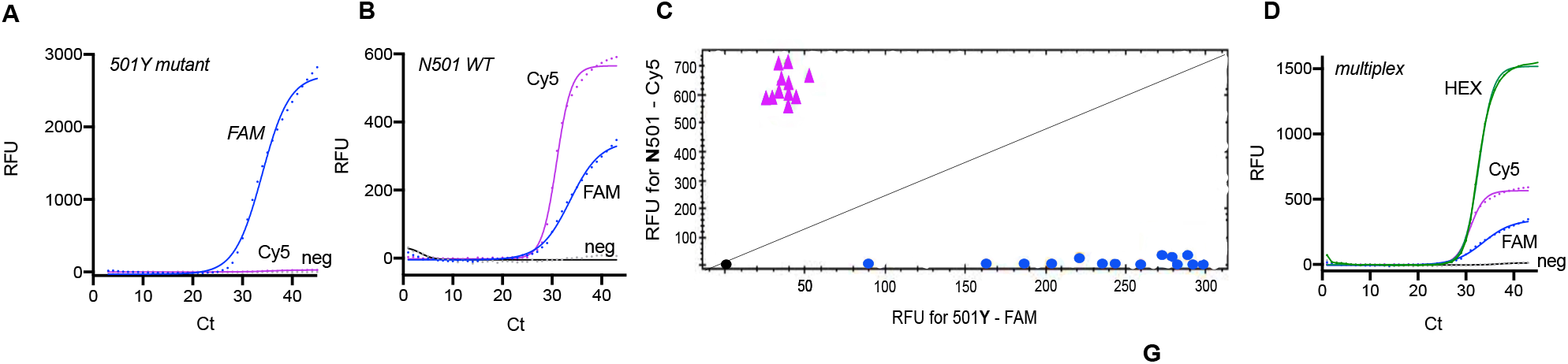
LNA-modified probes detecting the N501Y mutation. **A-B)**LNA probes detecting the 501Y mutation and N501 WT sequence respectively. **C)** Allelic discrimination analysis to differentiate between the 510Y mutation and N501WT sequence. **D)** Multiplexed PCR to detect the ΔH69/V70 mutation and the N501Y mutation.

## Acknowledgements

We would like to extend our gratitude to Susanne Lopez Rasmussen and Halenur A. Bayhan for their assistance and technical support. We would like to acknowledge the originating laboratories and WHO Collaborating Centre for Reference and Research on Influenza, The Francis Crick Institute, London, United Kingdom for providing reference virus material.

## Author contributions

Conceptualization: K.S, V.G., E.M., A.A.N., S.M., M.R., R.L., M.W.R., C.P., J.F., A.S.C, C.N. and A.F. Methodology: K.S., V.G., E.M., S.H.N., C.P., M.G.P.J, L.N. and DCGC. Investigation/Analysis: K.S., V.G., S.M.K., M.R., J.F. and A.S.C Visualization and data curation: K.S, V.G, E.M., A.A.N. and J.F. Supervision: A.F., A.S.C and C.P. Writing original draft: K.S. V.G. and A.F. Writing reviewing/editing: K.S., V.G., E.M., S.H.N., M.J., A.S.F., L.N., A.A.N., S.M.K., DCGC., S.M., M.R., R.L., M.W.R., C.P., J.F., A.S.C., C.N. and A.F. All authors critically revised the manuscript for important intellectual content and gave final approval for the submitted version.

